# Discriminating Progressive Supranuclear Palsy from Parkinson’s Disease using wearable technology and machine learning

**DOI:** 10.1101/19006866

**Authors:** Maarten De Vos, John Prince, Tim Buchanan, James J. FitzGerald, Chrystalina A. Antoniades

## Abstract

**Background:** Progressive supranuclear palsy (PSP), a neurodegenerative conditions may be difficult to discriminate clinically from idiopathic Parkinson’s disease (PD). It is critical that we are able to do this accurately and as early as possible in order that future disease modifying therapies for PSP may be deployed at a stage when they are likely to have maximal benefit. Analysis of gait and related tasks is one possible means of discrimination.

**Research Question:** Here we investigate a wearable sensor array coupled with machine learning approaches as a means of disease classification.

**Methods:** 21 participants with PSP, 20 with PD, and 39 healthy control (HC) subjects performed a two minute walk, static sway test, and timed up-and-go task, while wearing an array of six inertial measurement units. The data were analysed to determine what features discriminated PSP from PD and PSP from HC. Two machine learning algorithms were applied, Logistic Regression (LR) and Random Forest (RF).

**Results:** 17 features were identified in the combined dataset that contained independent information. The RF classifier outperformed the LR classifier, and allowed discrimination of PSP from PD with 86% sensitivity and 90% specificity, and PSP from HC with 90% sensitivity and 97% specificity. Using data from the single lumbar sensor only resulted in only a modest reduction in classification accuracy, which could be restored using 3 sensors (lumbar, right arm and foot). However for maximum specificity the full six sensor array was needed.

**Significance:** A wearable sensor array coupled with machine learning methods can accurately discriminate PSP from PD. Choice of array complexity depends on context; for diagnostic purposes a high specificity is needed suggesting the more complete array is advantageous, while for subsequent disease tracking a simpler system may suffice.

## Introduction

Progressive supranuclear palsy (PSP) (1, 2) is an atypical parkinsonian disorder that can sometimes be difficult to discriminate clinically from the much commoner Parkinson’s disease (3). It is characterized by vertical supranuclear gaze palsy, postural instability and axial rigidity as well as mild cognitive impairment (1) (4-6). Recently, the Movement Disorders Society (MDS) study group has proposed a set of diagnostic criteria for PSP, with the aim of improving the detection of the disease in clinical practice and research (7).

Comparative studies using accelerometers (8) have shown many shared gait abnormalities (9) (10-12) including decreases in velocity, step length, cadence, and mean acceleration. Some differences have also been found, including lower vertical displacement and higher acceleration in PSP. In this paper we examine the ability of a wearable array of inertial measurement units (IMUs), coupled with machine learning algorithms, to distinguish PSP patients from PD patients and from healthy controls.

There are a large and growing number of wearable technologies (13, 14) available for characterizing and measuring the motor features of neurodegenerative diseases(15-17).

When deciding what measurement technology to use there are two major issues that must be considered. One is the setting for the measurement, which can be broadly divided into laboratory measurements or ambulatory measurements outside the laboratory (e.g. at home). The former will permit closely controlled and supervised conditions giving the best chance of obtaining a high quality standardized dataset, and is well suited to tasks like detailed measurement of individual components of the gait cycle. However there is growing interest in longer term ambulatory measurement because snapshot laboratory recordings may not give an accurate overall picture, due to many factors such as variability of symptoms by time of day, inconsistent timing of medication, or the stress of being tested in a hospital environment. Laboratory measurements are also unlikely to capture infrequent but important features such as falls or near-falls, and a ‘real world’ environment may possibly better elicit features that are of direct clinical relevance to the patient.

The second issue is the choice of device. In particular, there is a conflict between maximising data and minimising the effort involved in obtaining it. At one end of the spectrum, it is possible to have body worn arrays comprising numerous inertial measurement units attached to all four limbs and the trunk, streaming tens of megabytes per minute. At the opposite extreme, some investigators are trying to develop systems based on single sensors, even those built into consumer smartphones, so that all that is required is an app running in the background. There is no doubt that the complex systems will generate much more complete information; the question is can the simpler systems answer the same diagnostic and measurement questions acceptably well, despite having a comparatively meagre dataset?

The two issues are clearly linked. Applying a complex system in a laboratory setting is straightforward and is logical given the intent of obtaining detailed data in a concentrated period of time. Using a complex system at home is another matter: donning and doffing may not be easy, and the multiple sensors may get in the way of everyday activities. There is a premium on using as simple a system as is compatible with obtaining sufficient data to be useful.

Our aim in this study is to answer two questions. Firstly, we will use a complex laboratory system with sensors on each limb and the trunk, to record data during commonly applied gait-related tasks. We will explore how well the data can distinguish PSP patients from PD patients and from healthy control (HC) participants, and what the most essential distinguishing features within the dataset are. Secondly, we will analyse which sensors/body locations are necessary to acquire this data. The aim is to determine how far the sensor array can be simplified while still yielding satisfactory results. It is hoped that this will help maximise compliance with future study protocols while avoiding collecting an inadequate dataset due to oversimplification.

## Materials & Methods

### Participants

The participants in this study were recruited as part of the Oxford study of Quantification in Parkinsonism (OxQUIP); a large clinical observational study being conducted at the John Radcliffe Hospital, Oxford. We recruited 20 participants with PD, 21 participants with PSP, and 39 healthy control participants. All healthy control participants were spouses of the PD or PSP participants. The demographics of the participants are summarised in Table 1.

**Table 1.**
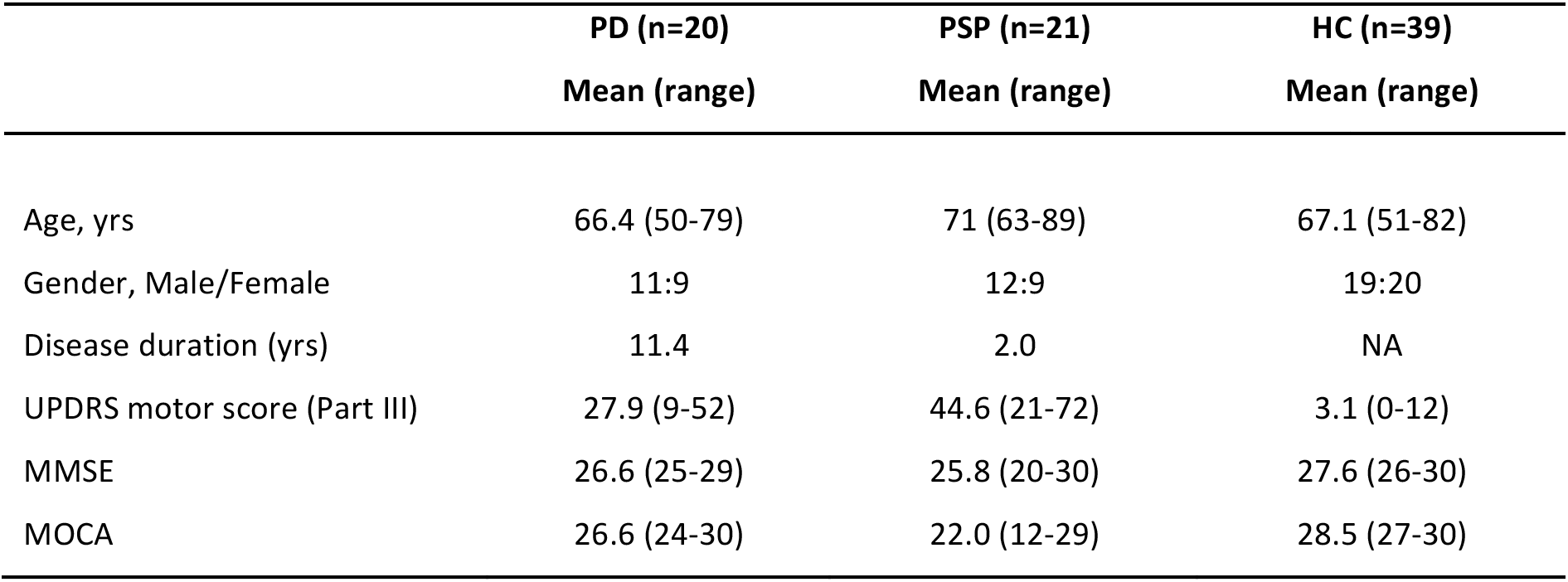
Demographics, clinical characteristics and cognitive scores in the 3 groups. PD = Parkinson’s disease, HC = healthy controls and PSP = Progressive Supranuclear Palsy. UPDRS = Unified Parkinson’s Disease Rating Scale, MMSE = Mini Mental state examination.

### Sensor array and software

Participants wore a system consisting of six synchronized inertial measurement units (IMUs) (Opal^™^, APDM, Portland, USA) that wirelessly transmitted data to software (Mobility Lab^™^, APDM) running on a nearby laptop. The IMUs were positioned over the lumbar spine, sternum, left and right wrists, and left and right feet. All sensors provided tri-axial accelerometer, tri-axial gyroscope, and tri-axial magnetometer signals at a frequency of 100Hz. Participants performed three tasks: two minute walk, sway test, and timed up-and-go (TUG). Using the waveforms from all sensors, the Mobility Lab software automatically extracts a range of clinical features specific to the three tasks. A full list of the task features are provided in figure 2. The full feature set included 109 parameters from analysis of the gait task, 33 from the sway test, and 14 from the TUG.

**Figure 1:**
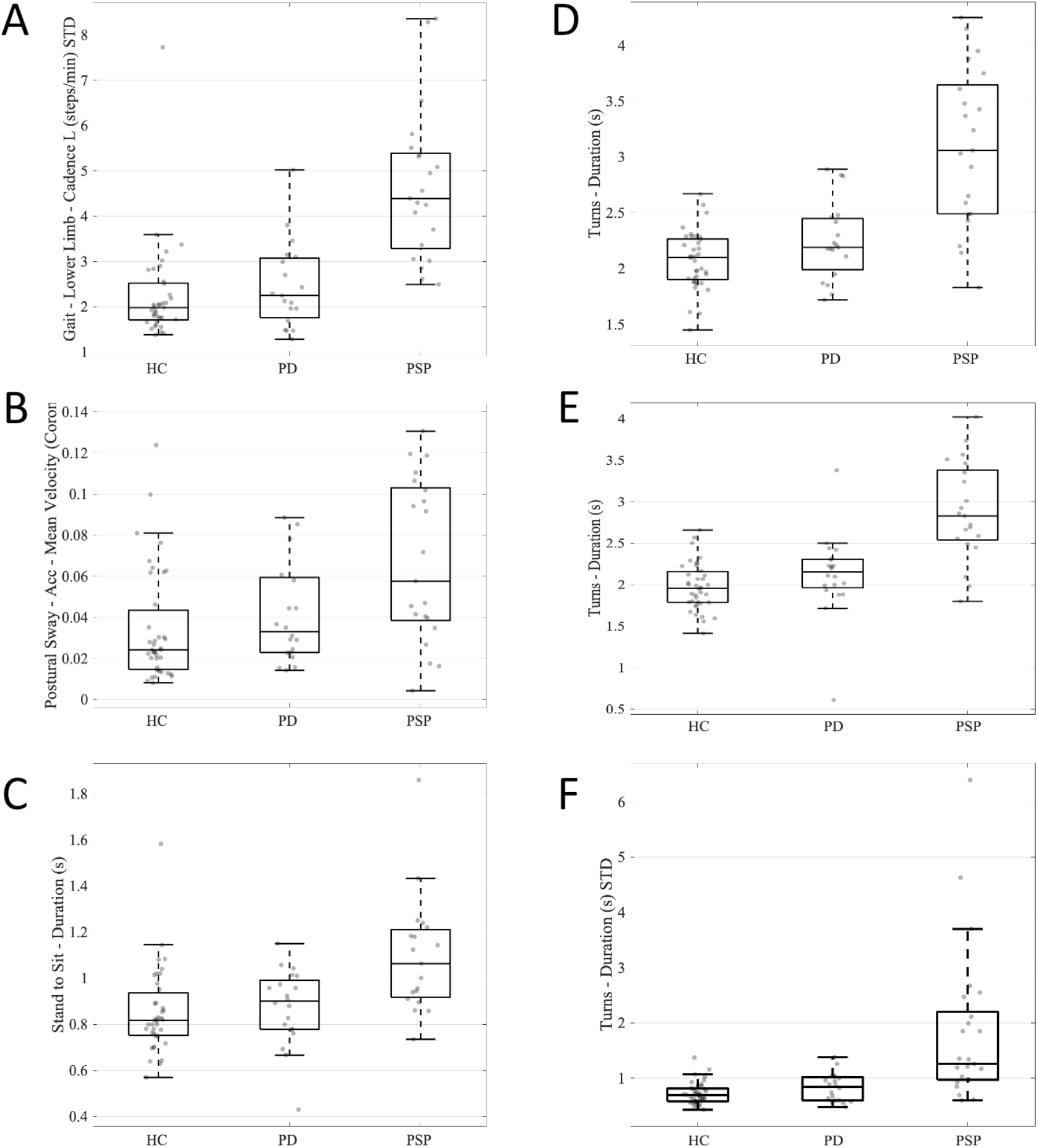
Examples of gait parameters that distinguish PSP from PD and HC. A: Gait cadence; B: Mean postural sway velocity in the coronal plane during the sway test; C: Mean time taken to sit from standing during the timed up-and-go (TUG) task; D: Mean time taken to turn during the gait task; E: Mean time taken to turn during the TUG task; F: Standard deviation of time taken to turn during the gait task. Note that the large number of parameters generated by the three tasks have considerable redundancy so that some parameters that are significantly different between the conditions individually are not in the LASSO output.

**Figure 2.**
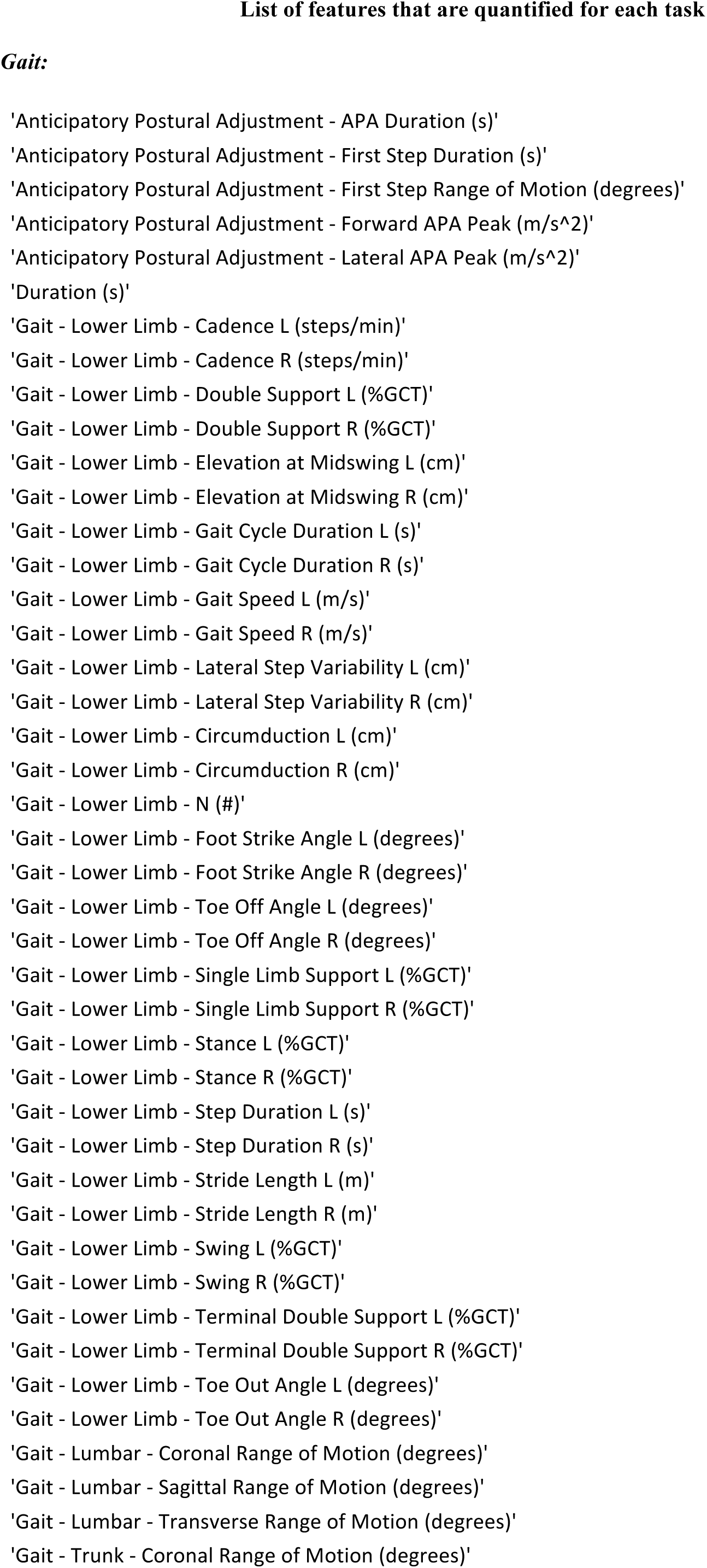
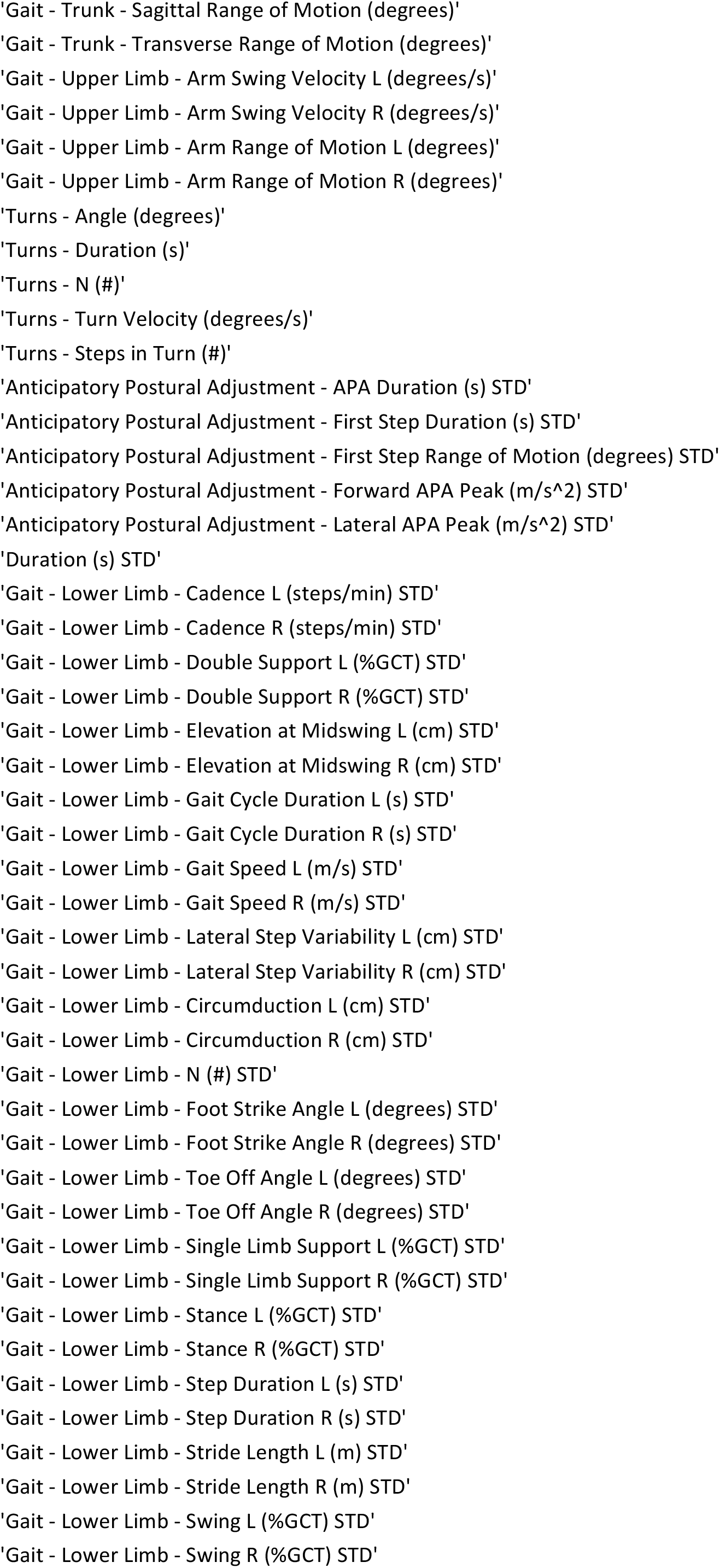
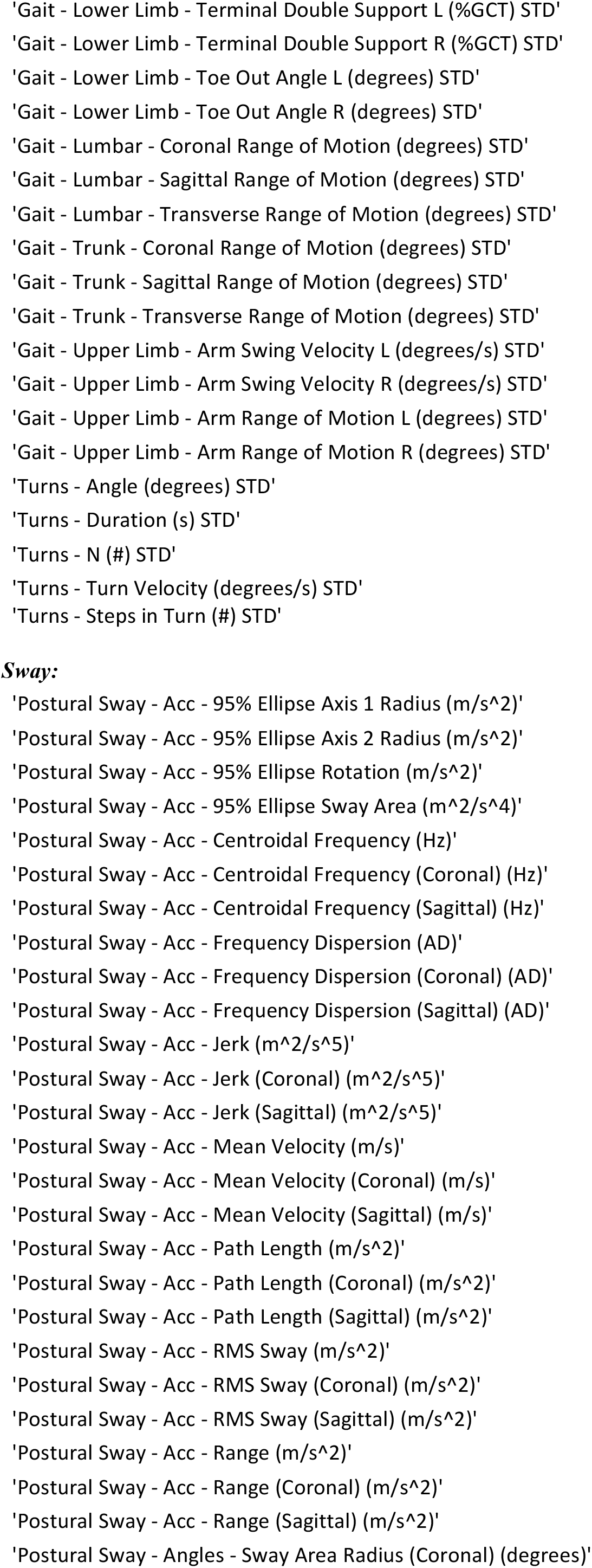
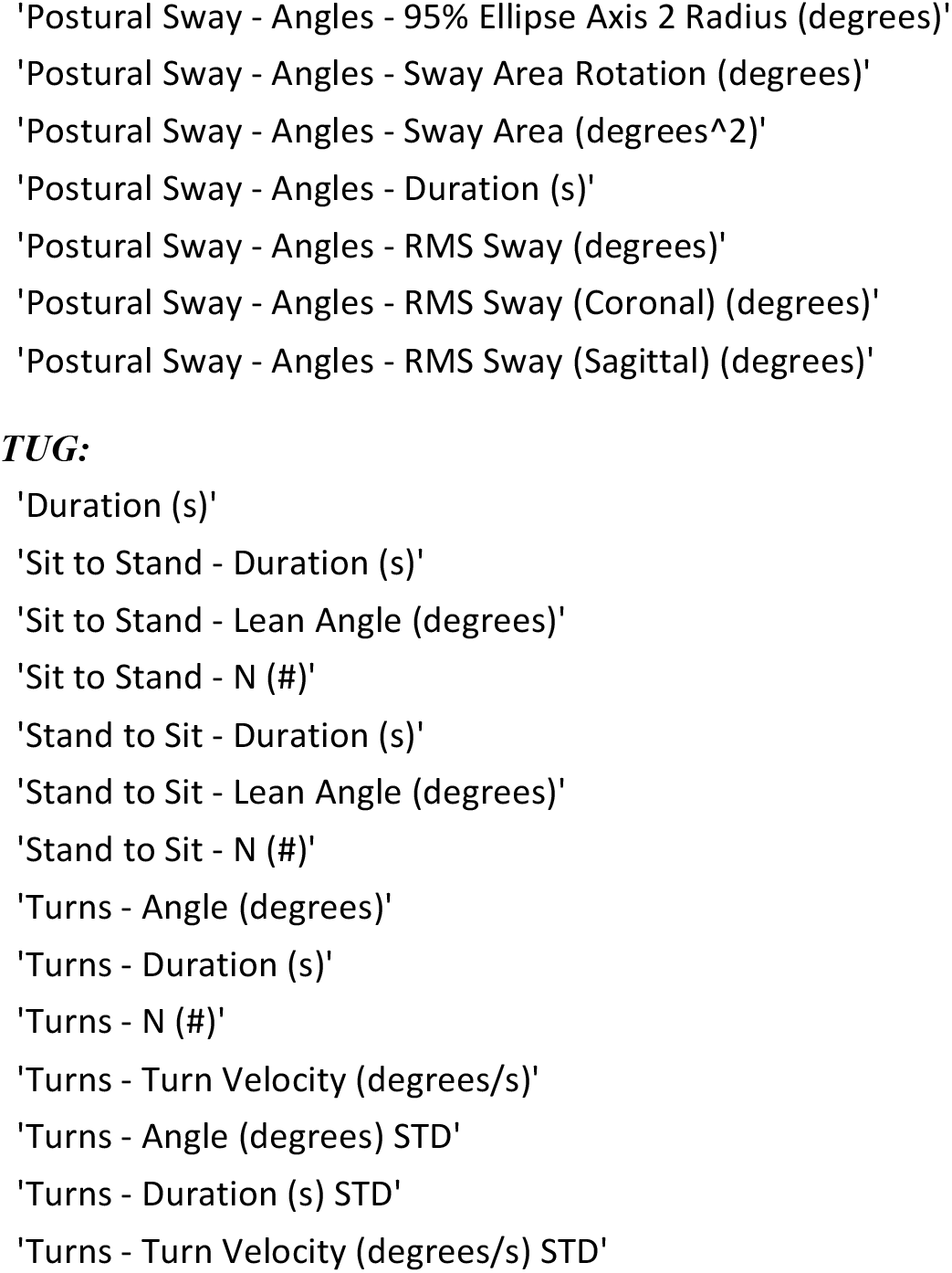
A full list of task features.

Any participants taking medication were recorded in the ‘ON medication’ state.

### Tasks

All participants completed a two minute walk on the same straight and level surface to record their gait. Next, to measure sway, participants were asked to stand upright and as still as possible for thirty seconds with their eyes closed on a firm surface. A wooden template was placed on the floor so as to ensure all participants’ feet were the same distance apart for the test. The TUG was then repeated three times. Each repetition entailed the participant starting in a sitting position for three seconds, followed by standing up when instructed, walking forward 3 metres, performing a 180 degree turn, walking back to the chair and sitting back down.

### Hypothesis Testing

The first aspect of this analysis aims to identify features that demonstrate statistically significant differences between the disease groups. Specifically, we aim to determine whether the PSP group shows a difference to HC and PD participants in a feature. Each participant contributes one instance in each of the three tasks. For each task type (gait, sway, and TUG), independent t-tests are used to inspect for statistical significance between the three disease groups for all features using a significance level of 0.05.

### Disease Classification

In order to assess to what extent PSP can be automatically discriminated from PD and HC subjects based on the measured features, we performed automated classification with a machine-learning algorithm. For each test, repeated 10-fold cross validation is performed based on the full feature set from each test separately. This means that the full dataset is split in randomly into 10 subsets, where 9 subsets are used for training the machine learning model and the remaining subset is used as an independent validation set. This training and validation is repeated 10 times where each time a different independent validation set is used. Within each repeat of cross validation, the cross sectional baseline subset undergoes minority class balancing prior to being assigned to a fold. Within each fold, the training and validation sets undergo zero-mean unit-variance normalisation with respect to the mean and standard deviation of the training set. Due to the likelihood that many of the features within each test are correlated (e.g. Gait Speed Left and Gait Speed Right), feature selection (Least Absolute Shrinkage and Selection Operator, LASSO) is performed on each training set, with the selected features subsequently being extracted from the validation set. Two classifiers were used. A Logistic Regression (LR) classifier was used due to its popularity in the clinical field. A Random Forest (RF) classifier was also used, without feature reduction as Random Forest classifiers are known to deal well with a high dimensional feature set. We report the accuracy, sensitivity, and specificity for each task separately and all tasks combined.

### Reduced sensor set

In order to assess the necessity or not of using a large sensor set, we took the features selected by LASSO for all tasks combined and assessed from which sensor those features are computed. We then repeated the classification procedure using only those features extracted from the lumbar sensor, because this sensor alone accounted for a majority of the features identified by LASSO. We further repeated the classification using data obtainable from the lumbar sensor plus the right arm and right leg sensors, and reported the same performance metrics.

## Results

### Participants

The demographics, clinical characteristics, and rating scale scores of the participants for each group are shown in table 1.

Figure 1 shows the distributions of some of the feature values extracted from the three tasks that were found to differ significantly between PSP and the two other groups. The box plots show the range of values for each of these features in each participant group, with individual data points overlaid in grey. Note that there is considerable redundancy amongst the large number of features, and not all measurements that were found to be significant individually emerged from the LASSO as independent predictors. For example in the sway test, mean coronal sway velocity was individually significant, but this did not feature in the LASSO result.

Table 2 shows the disease classification accuracy under a number of conditions. The upper three rows of the table show the accuracy, sensitivity, and specificity of discrimination between PSP and PD and between PSP and HC, for each of the three tasks separately as indicated in the left column. It can be seen that of the three tasks, sway is much less effective than the other two at discriminating between PSP and PD. The fourth row of table 2 gives the results of analysing the combined feature set from all three tasks; this performs better than any of the 3 tasks individually.

**Table 2.** Classification results when automatically classifying different patients based on their gait, sway, and TUG signatures using the entire six-sensor array (first three rows of table). Sway is the least informative test. Classification accuracy is improved when the feature sets from all three tasks are merged (fourth row). The Random Forest (RF) classifier performs better than the logistic regression (LR) classifier. Using data from the lumbar sensor alone (row 5) reduces accuracy modestly, but this can be recovered by adding back in arm and foot sensors from one side of the body (last row of table). The high specificity of PSP versus PD classification obtained with RF (90%) is only seen when using the 6 sensor array.

| Task | Sensors used | Comparison | Logistic Regression |  |  | Random Forest |  |  |
| --- | --- | --- | --- | --- | --- | --- | --- | --- |
|  |  |  | Acc (%) | Sens (%) | Spec (%) | Acc (%) | Sens (%) | Spec (%) |
| Gait | Lumbar, sternal, both arms, both feet | PSP vs HC | 87 | 86 | 87 | 91 | 85 | 94 |
|  |  | PSP vs PD | 80 | 81 | 75 | 83 | 81 | 85 |
| Sway | Lumbar, sternal, both arms, both feet | PSP vs HC | 68 | 52 | 77 | 82 | 67 | 90 |
|  |  | PSP vs PD | 63 | 66 | 60 | 63 | 67 | 60 |
| TUG | Lumbar, sternal, both arms, both feet | PSP vs HC | 90 | 81 | 97 | 92 | 86 | 95 |
|  |  | PSP vs PD | 70 | 71 | 70 | 83 | 86 | 80 |
| Combined | Lumbar, sternal, both arms, both feet | PSP vs HC | 93 | 86 | 97 | 95 | 90 | 97 |
|  |  | PSP vs PD | 80 | 85 | 75 | 88 | 86 | 90 |
| Combined | Lumbar only | PSP vs HC | 93 | 85 | 97 | 95 | 90 | 97 |
|  |  | PSP vs PD | 78 | 76 | 80 | 85 | 86 | 85 |
| Combined | Lumbar, right arm, right foot | PSP vs HC | 93 | 89 | 96 | 93 | 90 | 97 |
|  |  | PSP vs PD | 80 | 85 | 75 | 88 | 90 | 85 |

It is clear that the RF classifier performs better than the LR classifier overall. When distinguishing PSP from PD the sensitivity, specificity, and accuracy of RF was as good as or better than LR in every condition in the table. The greatest difference was 15 percentage points in specificity using the combined tasks and full sensor set (75% with LR versus 90% with RF).

Table 3 lists the features that emerged from the LASSO analysis of the combined tasks as the most important discriminators between the groups, with their weightings and which sensor in the array the data are obtained from in each case. Notably, the lumbar sensor provides the data for ten of the seventeen features in this list, compared to just one feature that is dependent on the sternal sensor. This prompts one to ask what the results would be if the lumbar sensor were used alone. The answer (table 2 row 5) is that using only one sensor in the lumbar position reduces the accuracy of both classifiers modestly, from 80% to 78% with LR and from 88% to 85% with RF.

**Table 3.** Key parameters emerging from LASSO analysis discriminating PSP from PD and HC subjects, together with their relative importance (as reflected by the weights). The ‘sensor’ column explains which of the sensors in the array is responsible for collecting the data described, and the activity column on the left shows during which test the feature has been collected. The three columns on the right show how much of the feature set is available depending on the extent of sensor array used.

| Activity | Feature | Weighting | Sensor | All sensors | Lumbar only | Lumbar, arm, leg |
| --- | --- | --- | --- | --- | --- | --- |
| TUG | 'Turns - Duration (s)' | 0.11328 | Lumbar | X | X | X |
| XX | 'Gait - Lower Limb - Cadence L (steps/min) STD' | 0.09398 | Left Leg | X |  | X |
| TUG | 'Turns - Angle (degrees) STD' | 0.06248 | Lumbar | X | X | X |
| TUG | 'Sit to Stand - Duration (s)' | 0.04518 | Lumbar | X | X | X |
| Gait | 'Gait - Trunk - Sagittal Range of Motion (degrees) STD' | -0.03971 | Trunk | X |  |  |
| Gait | 'Gait - Lower Limb - Toe Off Angle R (degrees) STD' | 0.03182 | Right Leg | X |  | X |
| TUG | 'Turns - Duration (s) STD' | 0.03071 | Lumbar | X | X | X |
| TUG | 'Stand to Sit - Lean Angle (degrees)' | -0.03009 | Lumbar | X | X | X |
| TUG | 'Turns - Angle (degrees)' | -0.02602 | Lumbar | X | X | X |
| TUG | 'Duration (s)' | 0.01957 | Lumbar | X | X | X |
| Gait | 'Gait - Upper Limb - Arm Swing Velocity R (degrees/s)' | -0.01876 | Right Arm | X |  | X |
| Gait | 'Gait - Upper Limb - Arm Range of Motion R (degrees)' | -0.01631 | Right Arm | X |  | X |
| Gait | 'Turns - Steps in Turn (#) STD' | 0.01618 | Lumbar | X | X | X |
| Sway | 'Postural Sway - Acc - Frequency Dispersion (AD)' | 0.01395 | Lumbar | X | X | X |
| Gait | 'Gait - Upper Limb - Arm Range of Motion L (degrees)' | -0.00735 | Left Arm | X |  | X |
| Gait | 'Gait - Lower Limb - Toe Out Angle L (degrees)' | 0.00101 | Left Leg | X |  | X |
| Gait | 'Turns - Duration (s)' | 0.00025 | Lumbar | X | X | X |

We then explored the effect of adding one arm and one leg sensor to the lumbar sensor, making a three sensor array including lumbar, right arm and right foot. Where LASSO had identified left sided parameter we substituted the equivalent parameter from the opposite side, making the assumption that, for example, cadence measured from either leg would report a similar value. Using this intermediate size network, the accuracy with both classifiers was the same as for the full six sensor network (table 2 row 6). Notably however, the high specificity (90%) observed with the 6 sensor network was reduced to 85% with the lumbar sensor alone, and this was not recovered using 3 sensors.

## Discussion

We have shown that it is possible with a body-worn IMU array and machine learning methods to differentiate PSP from PD and PSP from control, with a high degree of accuracy. In order to ensure the robustness of these findings we used separate training and validation datasets for the analysis, and to provide validation we used two different classification algorithms. Overall, the Random Forest classifier performed better than the Logistic Regression classifier. LR is probably the most commonly used classifier in biomedical research and therefore the apparent superiority of RF is an important finding.

A recent review found 78 published studies of IMU based gait analysis, but only 16% of these used more than one sensor (18). We found just two previous studies looking at gait in PSP. One of these used a single IMU in the lumbar region (8), and detected only a difference in vertical displacement during gait between PSP and PD. The other used two sensors, one attached to each foot (19), and identified significant differences in gait speed and cadence. In both cases the information obtained is far less detailed than that obtained from multiple sensors in the present study.

As expected there is a high degree of redundancy in the measured features within and across the three tasks used. This is well illustrated by the fact that, for example, the mean velocity of postural sway in the coronal plane was significantly and substantially different between PSP and the other groups, yet did not occur as an independent predictor in the feature set selected by the LASSO analysis. Indeed only one sway variable was found to contain independent information.

From the point of view of overall classification accuracy, it may be possible to simplify the sensor array. Of the 17 key parameters identified by the LASSO analysis, 10 could be obtained from just the lumbar sensor. In distinguishing PSP from HC, the accuracy, sensitivity, and specificity using just this sensor was virtually identical to that obtained using the full sensor array. Using just the lumbar sensor did degrade performance in terms of test accuracy when differentiating PSP from PD, but only modestly (for the RF classifier there was a test accuracy of 88% with six sensors versus 85% with the lumbar sensor only). Using an intermediate 3 sensor array (lumbar, right arm, right foot) gave a test accuracy as good as the full six sensor array.

In some respects however the most important parameter is specificity, because a higher specificity increases positive predictive value (PPV), which is critical if PSP specific treatments are to be initiated based on the result. The specificity of 90% using the 6 sensor array and the RF classifier is remarkably good. Previous biomarker studies using diverse approaches have not yielded specificities as high as this. For example, MRI morphometry of the brainstem gave a specificity of 85%(20), while a meta-analysis of studies of probably the most promising CSF biomarker, neurofilament light chain, gave a specificity of 81%(21). Because of the low prevalence of PSP amongst patients presenting with parkinsonism, even the higher 85% figure translated into a PPV of just 57% (20). The improvement in PPV between a test with 85% and 90% specificity is substantial. In moving from the full 6 sensors to just the lumbar sensor, the 90% specificity of our RF classifier fell to 85%, an important reduction in efficacy. Performance was not restored by adding in two more sensors.

It should be noted that the job that the classifiers were asked to do in this study is considerably easier than the applications to which we hope the approach will ultimately extend. It is no surprise to see classification accuracies in excess of 90% when discriminating PSP patients from controls. The fact that the classification accuracy between PSP and PD was not far behind (88% with RF) may seem impressive, but we began with groups of clear-cut PSP and PD, who would be readily distinguishable clinically by an experienced movement disorders neurologist. For the technique to be most useful, it must do something that such a person finds challenging. The performance differential between more complete and simpler sensor sets may well widen with more difficult questions and will need to be carefully evaluated in each situation.

The most obvious next step in testing this approach would be to evaluate its effectiveness in the differential diagnosis of these conditions at an earlier stage, when there is still substantial clinical diagnostic uncertainty. This will require the capture of a large incident cohort of parkinsonian patients, in order that it turns out eventually to contain sufficient numbers of cases that manifest as PSP, rather than the far commoner PD. The other task that could benefit from the machine learning approach is the related one of disease severity quantification and progression tracking. This is an area of great interest because objective numerical biomarkers capable of doing this in PSP (and PD) are lacking, yet very much needed for trials of potential disease modifying drugs.

In conclusion, the answer to the question of whether the sensor network can be simplified depends on context and the exact parameters we are interested in. It may be that the more complex sensor array is better suited to the diagnostic setting, where specificity is paramount, but then a simpler arrangement can be used for quantification and tracking once the diagnosis is secure.

## Data Availability

Anonymised data will be shared by request from any qualified investigator.

## ACKNOWLEDGMENTS

We thank the patients and their families for their endless support during the OxQUIP study.

## AUTHORS’ ROLES

Research project: Conception, organization, execution CAA, JFF

Statistical analysis: Design, execution, review & critique MDV, JP, JJF and CAA

Manuscript: Drafting, review & critique MDV, JP, TB, JJF and CAA

